# Evaluation of the diagnostic accuracy and analytical sensitivity of the novel Xpert® Mpox (Cepheid®) and STANDARD™ M10 MPX/OPX (SD Biosensor) molecular point-of-care assays for the detection of Mpox virus in skin lesion swabs and upper-respiratory swab samples

**DOI:** 10.1101/2024.09.09.24313234

**Authors:** Alessandra Romero-Ramirez, Anushri Somasundaran, Konstantina Kontogianni, Jacob Parkes, Yusra Hussain, Susan Gould, Christopher T Williams, Dominic Wooding, Richard Body, Hayley E Hardwick, J Kenneth Baillie, Jake Dunning, Malcom G Semple, CONDOR steering group, ISARIC CCP UK investigators, Tom E Fletcher, Thomas Edwards, Devy Emperador, Ana I Cubas-Atienzar

## Abstract

**Objectives:** Evaluation of diagnostic accuracy of two point-of-care (POC) molecular diagnostic tests for the detection of monkeypox virus (MPXV): Xpert® Mpox (Cepheid, Inc., USA) and STANDARD™ M10 MPX/OPX (SD Biosensor, Inc., Korea).

**Methods:** Diagnostic accuracy of both platforms was evaluated using 53 upper-respiratory swabs (URS) and 32 skin lesions swabs (SS) collected from mpox and COVID-19 patients in the UK against the Sansure (Sansure Biotech Inc.) and the CDC reference qPCR tests. The analytical sensitivity of both platforms was assessed using a viral isolate from the lineage II, B.1.

**Results:** The limit of detection was 1×10^1^ pfu/ml for both tests. The overall sensitivity and specificity of the Xpert® Mpox was 97.67% [95% CI 87.71–99.94%] and 88.57% [95% CI 73.26–96.80%] and 97.44% [95% CI 86.52–99.94%] and 74.42% [95% CI 58.83–86.48%] comparing the Sansure and CDC qPCR, respectively and for the M10 MPX/OPX was 87.80% [95% CI 73.80–95.92%] and 76.60% [95% CI 61.97–87.70%] and 94.29% [95% CI 80.84–99.30%] and 86.67% [95% CI 73.21–94.95%] with the Sansure and CDC qPCR.

**Conclusion:** The Xpert® Mpox had good diagnostic accuracy for both sample types while the M10 MPX/OPX clinical accuracy was deficient with URS. Our data supports the use of URS during the first 3 days of symptoms onset for mpox diagnosis.

**Highlights:**

- The Xpert® Mpox detected Monkeypox virus (MPXV) DNA in more samples than the M10 MPX/OPX, CDC qPCR and Sansure qPCR suggesting higher sensitivity at lower viral loads.
- Xpert® Mpox fulfilled the performance requirements recommended in the World Health Organisation (WHO) target product profile (TPP) using upper-respiratory swabs (URS) and skin lesion swabs (SS) but M10 MPX/OPX performance was only met when using SS.
- MPXV DNA was reliably detectable in SS up to 5 days after onset of symptoms. with all PCR tests
- The use of URS for mpox diagnosis is not recommended for use more than 3 days after onset of symptoms.

## Introduction

The highly infectious monkeypox virus (MPXV) is a double-stranded DNA virus belonging to the *Orthopoxviridae* family, which includes vaccinia, cowpox, and variola viruses^1^. Orthopoxviruses are large viruses with a size range from 140-450 nanometers and a genome that contains over 200 genes^2^. MPXV was identified in 1958 in captive cynomolgus macaques (*Macaca fascicularis*) that were transported from Singapore to Denmark^3^ and in 1970, the first known case of MPXV infection in a human from the Democratic Republic of the Congo (DRC) was reported^4^. The WHO recommended “Mpox” as the preferred term for human disease caused by MPXV in November 2022^5^.

In May 2022, an mpox outbreak spread to over 110 countries with over 86,000 confirmed cases^6^. The number of infections during the 20^th^ century has already been surpassed by cases after the 2022 outbreak^7^. At that time, in the United Kingdom (UK), all clades of mpox were classified as a High Consequence Infectious Disease (HCID) and patients were looked after in specially designed HCID treatment facilities run by a nationwide network^8^. From August 2018 to September 2021, 7 mpox cases were identified in the UK and received treatment in HCID centres (4 imported cases and 3 secondary cases)^8^. The discovery of the first mpox case of the global outbreak was on May 7, 2022, a person who travelled from Nigeria^9^ and as of June 8, 2022, there were 336 laboratory-confirmed cases in the UK. Most of these cases were identified in men [99%], who were primarily residents of London [81%]^10^. For the first time, community transmission was reported in the UK, which was mainly through intimate person-to-person contact, often involving sexual activity and mostly unrelated to travel from endemic countries^11^.

Clinical manifestations of mpox infection include a vesiculopustular rash resembling that of smallpox, fevers, lymphadenopathy and a rash may affect palms and soles. Skin lesions may commence at the site of initially inoculation or exposure e.g. the anogenital region after transmission during sexual contact or at the site of a needlestick injury or bite^12^. According to the Centres for Disease Control and Prevention (CDC), the incubation period is up to 21 days following/after viral exposure and the rash appears 1-4 days after initial flu-like prodrome^13^. To confirm a clinical diagnosis, the World Health Organisation (WHO) advises testing for mpox as soon as possible in people who fit the suspected case definition^14^. Laboratory-based nucleic acid amplification tests (NAAT) are the primary method used for mpox diagnosis^15^. Laboratory-based PCR testing requires specialist equipment, up front DNA extraction, and skilled personnel to perform such tests. Many cases in low- and middle-income countries (LMIC) remain unreported due to a lack of decentralised diagnostic resources in the area, and issues with the current healthcare system and civil upheaval.

The increasing global cases of mpox following the 2022 outbreak brought to light the difficulties in meeting the increased and erratic demand of decentralised diagnostics for different virus prone to outbreaks. Another public health emergency of international concern (PHEIC) was declared by WHO on 14^th^ August 2024 given the significant increase in mpox cases which has the potential to spread beyond Africa^16^. This highlighted the urgent need for the rational development of rapid diagnostic methods for emerging pathogens such as for MPXV as a priority. As a result, several NAAT were developed to identify MPXV at the point-of-care (POC) since the 2022 outbreak^17,18^. POC NAAT offer higher sensitivity and specificity compared to antigen-based POC tests and are equal to laboratory-quality testing without the requirement for sophisticated laboratory facilities^19^, requiring less operational training and fewer sample preparation steps compared to lab-based PCR.

Prompt isolation and optimal clinical care are all dependent on an accurate diagnosis of MPXV infection. In this study, we evaluated the diagnostic accuracy of two new POC NAAT, Xpert® Mpox (Cepheid, Inc., Sunnyvale, CA, USA) and STANDARD™ M10 MPX/OPX (SD Biosensor, Inc., Suwon, Korea), for the detection of MPXV on skin lesion and upper-respiratory swabs.

## Methodology

### Study design

Skin lesion swabs (SS) (n=30) and upper-respiratory swab (URS) samples (n=23, [nasopharyngeal=1, oropharyngeal=22]) in universal transport media (UTM, RT-UTM Copan, Italy) from a cohort of 16 mpox patients enrolled at the Royal Liverpool University Hospital, Sheffield Teaching Hospital NHS Foundation Trust, and Royal London Hospital were used for this study. Patients were recruited during the last two outbreaks of mpox in the UK, 2018 and 2022. Patients were consented under the WHO ISARIC4 Comprehensive Clinical Characterisation Collaboration Protocol for severe emerging infections [ISRCTN66726260]^20^, ethical approval was obtained from the National Research Ethics Service and the Health Research Authority (IRAS ID:126600, REC 13/SC/0149). All mpox patients were diagnosed by sending samples to the UK Health Security Agency (UKHSA) for testing using qPCR. In addition to the samples from mpox positive patients, to fulfil with the minimum number of negative swab specimens for Mpox diagnostic evaluations recommended by the by the FDA^15^, a set of 32 leftover nasopharyngeal samples in UTM (RT-UTM Copan, Italy) from prior COVID-19 studies^19–22^ were used as mpox negative controls. These were collected under the Facilitating AcceLerated Clinical validation Of Novel diagnostics for COVID-19^19,23^ and ethical approval was obtained from the National Research Ethics Service and the Health Research Authority (IRAS ID:28422, REC: 20/WA/0169). All samples were aliquots stored at -80°C and thawed for the first time for this study. Samples were processed and tested at the Biosafety Level 3 (BSL3) Laboratories of the Liverpool School of Tropical Medicine (LSTM) as previously described^19^.

### MPXV PCR reference assays

The DNA was extracted from 200µL of UTM using the QiAamp96 Virus Qiacube HT kit (Qiagen, Germany). Two reference PCR tests were used, the commercially available CE-IVD Sansure qPCR kit (Monkeypox virus Nucleic Acid Diagnostic Kit, Sansure Biotech Inc.), and the CDC Monkeypox virus Generic Real-Time PCR Test^21^. Both lab-based PCR tests were used as reference tests as the CDC qPCR is widely used, the Sansure qPCR kit is CE-IVD marked and both have successfully demonstrated to detect MPXV clades I, IIa and IIb^22,23^. The PCRs were performed on the QuantStudio 5 (ThermoFisher, USA) following the manufacturer instructions (Sansure Biotech Inc.) and the CDC guidelines^21^. The CDC qPCR was performed using the QuantiFast Pathogen PCR kit (Qiagen, Germany).

### MPXV POC index NAAT

Two rapid molecular POC platforms which perform automated sample processing and qPCR to detect viral DNA were evaluated in the study: Xpert® Mpox and the STANDARD™ M10 MPX/OPX (M10 MPX/OPX hereinafter). The platforms were selected following an expression of interest launched by FIND (www.finddx.org) and a scoring process based on defined criteria. The evaluation of the platforms at LSTM was done in BSL3 laboratories.

The Xpert® Mpox assay is authorized for use under FDA Emergency Use Authorization (EUA) and provides semiquantitative detection and differentiation between MPXV clade II (two undisclosed targets) and non-variola *Orthopoxvirus* (target OPXV-E9L NVAR gene) DNA, respectively^24^. A Sample Processing Control (SPC), a Sample Adequacy Control (SAC), and a Probe Check Control (PCC, not included in the algorithm, used as quality control) are also included in the cartridge utilised by the GeneXpert® instrument^25^. The tests were performed according to the manufacturer’s instructions. Briefly, 300µL sample were transferred to the sample chamber of the Xpert^®^ Mpox test cartridge and loaded onto the GeneXpert^®^ Instrument System platform. The results were automatically interpreted by the GeneXpert^®^ System based on the Ct values results. A sample was called positive when it was positive for the 2 MPXV targets (OPXV, SAC, SPC could be either positive or negative); negative result when it was negative for MPXV and OPXV but positive for SAC and SPC; a positive result for non-variola OPXV when it was positive for the OPXV target, negative for MPXV (SAC and SPC can be either positive or negative); and invalid when it was negative for both viral targets and controls or when only one control was positive but both viral targets negative.

The M10 MPX/OPX assay is for Research Use Only and provides semiquantitative detection and differentiation between MPXV and OPXV DNA using E9L and G2R gene targets, respectively. The LOD as reported by the manufacturer is 100 copies/ml. The tests were performed according to manufacturer’s instructions. Briefly, 300µL of sample were transferred to the sample chamber of the cartridge and loaded onto the STANDARD™ M10 platform. After 1 hour, the results with the corresponding Ct values were displayed on the STANDARD™ M10 screen and the results were automatically interpreted. A sample was considered positive for MPXV when MPXV and OPVX targets were positive (IC can be either positive or negative), positive result for OPXV when MPXV was negative and OPXV was positive (IC can be either positive or negative), negative when only the IC was positive and invalid when all targets were negative or when only the MPXV target was positive.

### Analytical limit of detection of qPCR reference assays and POC index NAATs

A MPXV strain (Slovenia_MPXV-1_2022, isolate 2225/22 Slovenia ex Gran Canaria) from the lineage II, B.1 (European Virus Archive Global EVAg, Marseille, France) was cultured in Vero C1008 (ECACC 85020206) (Vero E6 cells) obtained from the European collection of authenticated cell cultures (ECACC) in Dulbecco’s Modified Eagle Medium (DMEM, Gibco, USA) plus 10% foetal bovine serum (FBS, Gibco, USA) and 1% Penicillin/Streptomycin solution (Gibco, USA) to generate the MPXV stock. Frozen aliquots of the fourth passage of the virus were quantified via plaque assay. The MPXV stock was used to investigate the limit of detection (LOD) of both Xpert® Mpox and M10 MPX/OPX assays. A fresh aliquot was serially diluted from 1.0x 10^4^ plaque forming units (pfu)/ml to 1.0 x 10^2^ pfu/ml using UTM media. Each dilution was tested in triplicate and UTM was used as negative control following previous work^26–29^. The LOD was defined as the lowest dilution where all the three replicates were positive. DNA from the serial dilutions was extracted using the QiAamp96 Virus Qiacube HT kit and viral copy numbers per mL (copies/mL) were calculated using a standard curve of quantified synthetic DNA (G2R gene) in the QuantStudio 5 tested using the CDC PCR. Synthetic DNA (Eurofins Genomics, UK) was re-hydrated in Tris-EDTA buffer and concentration quantified using Qubit^TM^ SSDNA Quantification Assay kit (ThermoFisher, USA). Standard curve was prepared using an eight 10-fold serial dilution series with 5 replicates per dilution.

### Statistical analysis

The sensitivity and specificity of the index tests were calculated with 95% confidence interval in comparison to both reference PCR assays, including stratification by cycle threshold (Ct) value. Prior the analysis, a normality test was performed using the Shapiro Wilk test (p<0.05). Differences between the Ct values (expressed as mean± standard deviation [SD]) in sample groups were assessed using the paired Student’s t-test. Differences in the frequency of MPXV detection by sampling date were analysed using Chi-squared test and Fisher’s exact test. Statistical significance was set for a p <0.05. The statistical analysis was performed using GraphPad Prism version 8.0.0, GraphPad Software (Boston, USA).

## Results

### Clinical evaluation

The demographic and clinical characteristics of the participants are shown in Table 1. The 16 positive patients with mpox were assessed at the three hospitals during the study period. All individuals were men (100%) with a mean age of 35.1 years (range 24-58 years). The median days from onset of symptoms was 8 with the most common symptoms being skin lesions (100%), skin rushes (87.5%) and fever (68,75%). The results shown in table 1 included only the mpox positive patients (n=16). The negative cohort (COVID-19 patients) was not included in Table 1 as these were from a population not suspected from MPXV infection.

**Table 1.** Clinical characteristics of mpox patients from UK used for the evaluation of both molecular platforms.

| Characteristic |  |
| --- | --- |
| Age [mean (min-max), N] | 35.1 (24-58), 16 |
| Gender [%M, (n/N)] | 100%; (16/16) |
| Days from symptom onset [median (Q1-Q3); N] | 8 (4.25 - 12.75); 16 |
| Days < 0–3 (n, %) | 1, 6.25% |
| Days 4–7 (n, %) | 6, 37.5% |
| Days 8+ (n, %) | 9, 56,25% |
| **Symptoms [% (n/N)]** |  |
| Skin lesions | 100% (16/16) |
| Skin rashes | 87.5% (14/16) |
| Fever | 68.75% (11/16) |
| Flu like symptoms | 25% (4/16) |
| Headache | 25% (4/16) |
| Sore throat | 25% (4/16) |
| Cough | 6.25% (1/16) |
| Diarrhoea | 6.25% (1/16) |
| Chest pain | 0% (0/16) |
| Abdominal pain | 0% (0/16) |
| Nausea | 0% (0/16) |
| Vomiting | 0% (0/16) |
| Painful Urination | 0% (0/16) |

Fourteen of the 23 URS and 25 of 30 SS collected from mpox positive patients were positive by the CDC qPCR. When using the Sansure qPCR, 1 further URS (4.3%) and 4 SS (13%) were positive. The mean Ct value when using the Sansure and CDC qPCR were 30.09 (**±** 5.70) and 27.54 (**±**5.87), respectively. The mean difference in Ct values between both tests had no significant difference for any of the sample types (p-value= 0.34). As expected, all 32 UTM samples collected from the COVID-19 cohort were negative for MPXV using both reference qPCR tests.

The overall clinical sensitivity and specificity for the Xpert® Mpox assay using both sample types were 97.67% [95% CI 87.71 – 99.94%] and 88.57% [95% CI 73.26 – 96.80%] with the Sansure qPCR and 97.44% [95% CI 86.52 – 99.94%] and 74.42% [95% CI 58.83 – 86.48%] comparing to the CDC qPCR.

The values by sample type are found in Tables 2 and 3. The overall percentage of agreement was 90.3% [95% CI 81.7– 95.7%] and 91.5% [95% CI 83.2– 96.5%], when using the Sansure qPCR and CDC qPCR. Three URS were invalid with Xpert® Mpox assay (5.45%, 3/55) and all of SS were valid. (Tables 2-3). The specificity was 83.78% [95% 78.20 - 100%] and 81.58% [95% CI 65.67 – 92.26%] for URS using Sansure and CDC PCR. Specificity could not be accurately calculated for SS due to the lack of negative specimens using the reference tests.

**Table 2.** Results and clinical sensitivity and specificity of the Xpert^®^ Mpox assay and M10 MPX/OPX using upper-respiratory samples from mpox (n=23) and COVID-19 patients (n=32).

|  | Results | Sansure qPCR |  |  | CDC qPCR |  |  |
| --- | --- | --- | --- | --- | --- | --- | --- |
|  |  | Positive | Negative | Total | Positive | Negative | Total |
| Xpert® Mpx | Positive | 15 | 6 | 21 | 14 | 7 | 21 |
|  | Negative | 0 | 31 | 31 | 0 | 31 | 31 |
|  | Total* | 15 | 37 | 52 | 14 | 38 | 52 |
|  | Clinical sensitivity (95% CI) | 100% (78.20% - 100%) |  |  | 100% (76.24% - 100%) |  |  |
|  | Ct <25 [95% CI, N] | 100% (15.81% - 100%), 2 |  |  | 100% (39.76% - 100%), 4 |  |  |
|  | Ct <35 [95% CI, N] | 100% (69.15% - 100%), 10 |  |  | 100% (69.15% - 100%), 10 |  |  |
|  | Ct <40 [95% CI, N] | 100% (78.20% - 100%), 15 |  |  | 100% (76.84% - 100%), 14 |  |  |
|  | Clinical specificity (95% CI) | 83.78% (67.99% - 93.81%) |  |  | 81.58% (65.67% - 92.26%) |  |  |
| M10 MPX/OPX | Positive | 10 | 2 | 12 | 10 | 2 | 12 |
|  | Negative | 4 | 36 | 40 | 2 | 38 | 40 |
|  | Total* | 14 | 38 | 52 | 12 | 40 | 52 |
|  | Clinical sensitivity (95% CI) | 71.43% (41.90% - 91.61%) |  |  | 71.43% (41.90% - 91.61%) |  |  |
|  | Ct <25 [95% CI, N] | 100% (15.81% - 100%), 2 |  |  | 100% (15.81% - 100%), 2 |  |  |
|  | Ct <35 [95% CI, N] | 100% (66.37% - 100%), 9 |  |  | 100% (66.37% - 100%), 9 |  |  |
|  | Ct <40 [95% CI, N] | 71.43% (41.90% - 91.61%), 10 |  |  | 100% (69.15% - 100%), 10 |  |  |
|  | Clinical specificity (95% CI) | 92.11% (78.62% - 98.34%) |  |  | 94.59% (81.81% - 99.34%) |  |  |

\*Three samples were invalid with Xpert® Mpx and M10 MPX/OPX.

**Table 3.** Results and clinical sensitivity and specificity of the Xpert^®^ Mpox assay and M10 MPX/OPX assays using skin lesion swabs (SS) (n=30).

|  | Results | Sansure qPCR |  |  | CDC qPCR |  |  |
| --- | --- | --- | --- | --- | --- | --- | --- |
|  |  | Positive | Negative | Total | Positive | Negative | Total |
| Xpert® Mpx | Positive | 27 | 2 | 29 | 24 | 4 | 28 |
|  | Negative | 1 | 0 | 1 | 1 | 1 | 2 |
|  | Total | 28 | 2 | 30 | 25 | 5 | 30 |
|  | Clinical sensitivity (95% CI) | 96.43% (81.65% - 99.91%) |  |  | 96% (79.65% - 99.90%) |  |  |
|  | Ct <25 [95% CI, N] | 100% (63.06% - 100%), 8 |  |  | 100% (73.54% - 100%), 12 |  |  |
|  | Ct <33 [95% CI, N] | 100% (63.03% - 100%), 17 |  |  | 100% (83.16% - 100%), 20 |  |  |
|  | Ct <40 [95% CI, N] | 96.43% (81.65% - 99.91%), 30 |  |  | 96% (79.65% - 99.90%), 30 |  |  |
|  | Clinical specificity (95% CI)* | NA |  |  | NA |  |  |
| M10 MPX/OPX | Positive | 26 | 1 | 27 | 23 | 4 | 27 |
|  | Negative | 1 | 0 | 1 | 0 | 1 | 1 |
|  | Total** | 27 | 1 | 28 | 23 | 5 | 28 |
|  | Clinical sensitivity (95% CI) | 96.30% (81.03% - 99.91%) |  |  | 100% (85.18% - 100%) |  |  |
|  | Ct <25 [95% CI, N] | 100% (69.15% - 100%), 10 |  |  | 100% (73.54% - 100%), 12 |  |  |
|  | Ct <33 [95% CI, N] | 100% (79.41% - 100%), 16 |  |  | 100% (82.35% - 100%), 19 |  |  |
|  | Ct <40 [95% CI, N] | 96.30% (81.03% - 99.91%), 28 |  |  | 100% (85.18% - 100%), 23 |  |  |
|  | Clinical specificity (95% CI)* | NA |  |  | NA |  |  |

\*Only one SS was negative using the CDC qPCR so specificity was not calculated for this sample type. \*\*Two SS were invalid with M10 MPX/OPX assay.

The overall sensitivity and specificity for the M10 MPX/OPX using both sample groups were 87.80% [95% CI 73.80 – 95.92%] and 76.60% [95% CI 61.97 – 87.70%] and 94.29% [95% CI 80.84 – 99.30%] and 86.67% [95% CI 73.21 – 94.95%] compared with the Sansure and CDC qPCR, respectively. The values by sample type are found in Tables 2 and 3. The overall percentage of agreement was 91.3% [95% CI 82.8 – 96.4%] and 95.0% [95% CI 87.7 – 98.6%] with the Sansure qPCR and CDC qPCR, respectively. The specificity was 92.11% (78.62% - 98.34%) and 94.59% [95% CI 81.81 - 99.34%] using URS compared to Sansure and CDC PCR, respectively. All SS were positive with both reference assays except for 1 sample using the CDC PCR, therefore specificity could not be accurately calculated for this sample type. Three URS (5.45%, 3/55) and 2 SS (6.66%, 2/30) were invalid using the M10 MPX/OPX.

The Ct values of paired samples were also compared and evaluated for each qPCR reference assay and POC index NAAT (Figures 1A-1D). Nine, 10, 10 and 7 URS and SS paired samples were positive for Sansure qPCR, CDC, Xpert Mpox and M10 MPX/OPX. No significant differences in Ct values were found between URS and SS sample groups when using the Sansure qPCR, CDC and M10 MPX/OPX (p-value=0.54, 0.73 and 0.37, respectively). The analysis of the paired samples using the Xpert^®^ Mpox assay showed higher Ct values in the URS group compared to the SS group (p-value=0.03) with mean Ct values of 30.58 (± 5.48) and 24.75 (±5.98) for URS and SS.

**Figure 1.**
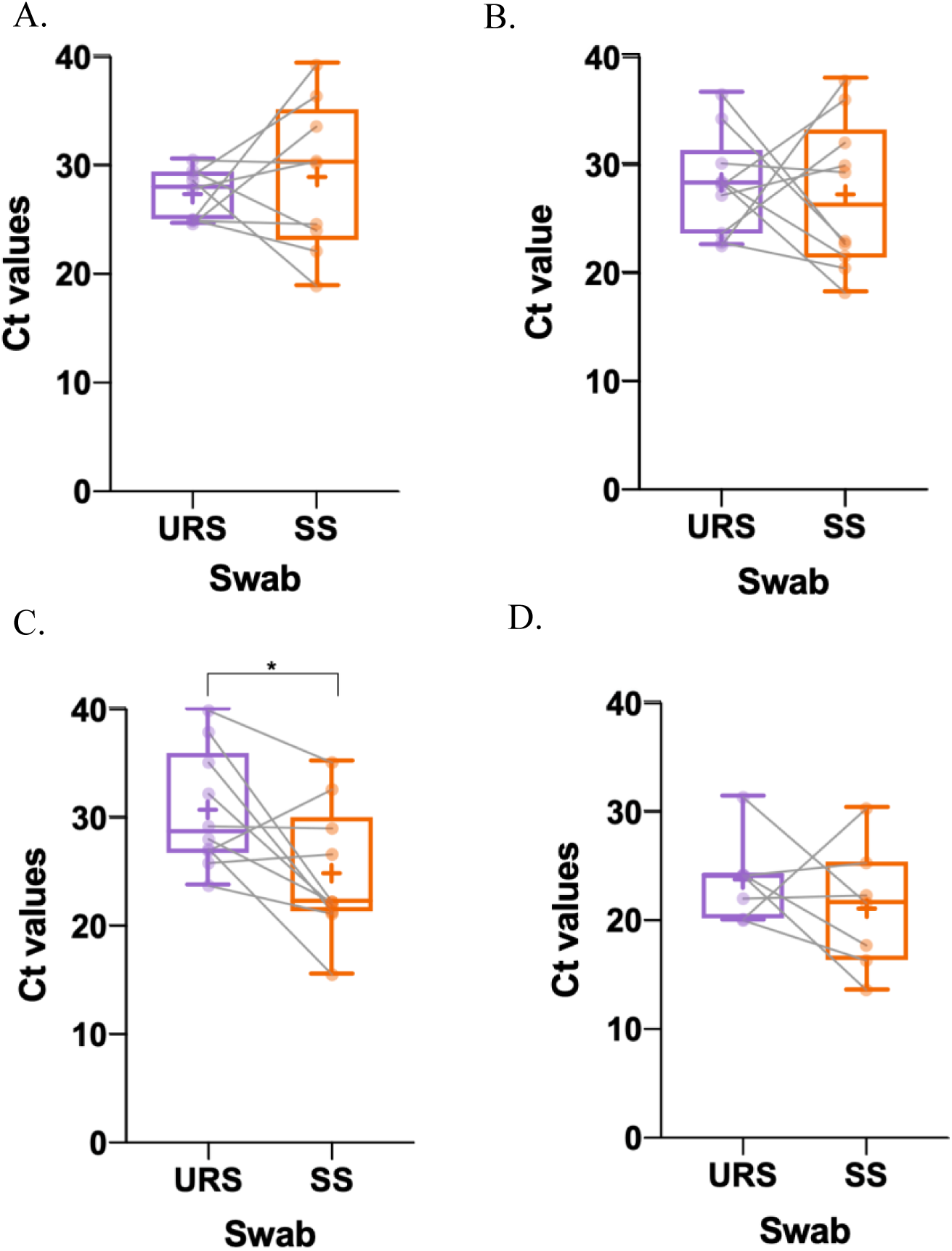
Boxplot of the Ct values from paired URS and SS tested by A. Sansure qPCR (n=9), B. CDC qPCR (n=10), C. Xpert^®^ Mpox (n=10) and D. M10 MPX/OPX (n=7). The whiskers show the maximum and minimum values and the vertical line the median. There was a significant difference (p-value <0.05) between paired URS and SS when evaluated with the Xpert^®^ Mpox assays with higher Ct values in the URS group.

Overall, the higher positivity rates for detecting MPXV DNA in clinical samples was the Xpert^®^ Mpox (n=50/53), followed by Sansure qPCR (n=44/53), CDC qPCR (n=41/53) and M10 MPX/OPX (n=37/53) and this difference was statistically significant for URS (p= 0.015) but not for SS (p= 0.692).

The number of MPXV positive samples depending on the sampling collection day from onset of symptoms was evaluated for all samples and all PCR tests (Figure 2). URS collected from MPXV patients more than 3 days after the symptom onset were less likely to have detectable levels of virus using all PCR assays used in the study (Sansure p = 0.017, CDC p = 0.033, Xpert^®^ Mpox p = 0.04 and M10 MPX/OPX p = 0.014). URS collected from MPXV patients more than 2 days after the symptom onset were less likely to present the virus to detectable levels by Sansure (p = 0.014) and M10 MPX/OPX (p = 0.007). This was not significant for the SS for any collection date (all p values > 0.05) except for M10 MPX/OPX among SS collected more than 5 days after onset of symptoms (p = 0.022).

**Figure 2:**
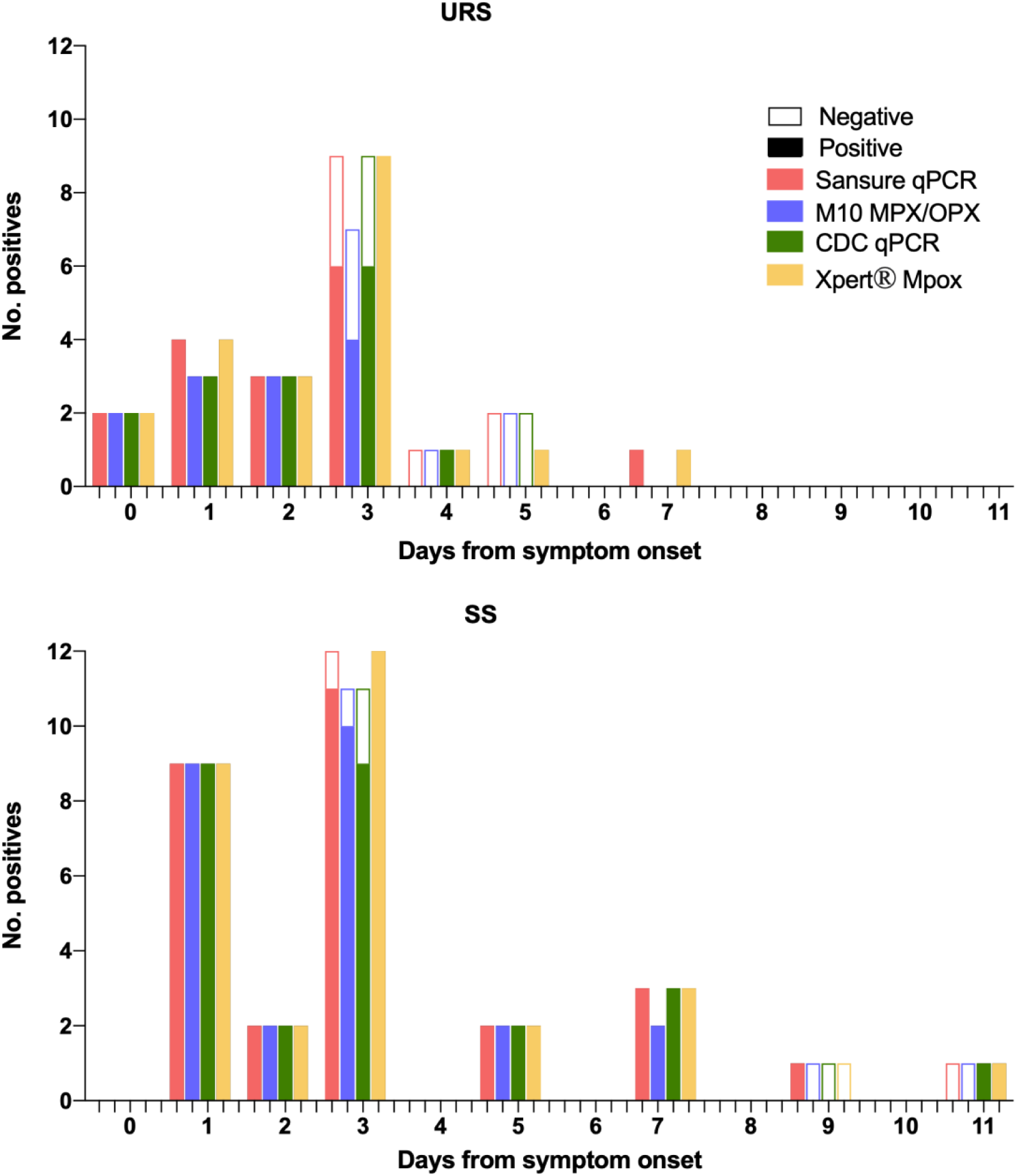
Number of positive samples according to sampling date from symptoms onset for SS and URS.

The Ct values as a proxy for viral loads were analysed by sampling day from onset of symptoms and higher Ct values were observed as the sampling day increased in URS for Sansure qPCR (p = 0.0093, r=0.58 95% 0.12-0.84) CDC qPCR (p = 0.0444, r=0.45 95% -0.08-0.8) and Xpert^®^ Mpox (p = 0.0024, r=0.59 95% 0.21-0.81) but not for M10 (p = 0.1752, r=0.30 95% -0.33-0.74). No correlation was observed between viral loads and sampling date in SS (Figure 3).

**Figure 3:**
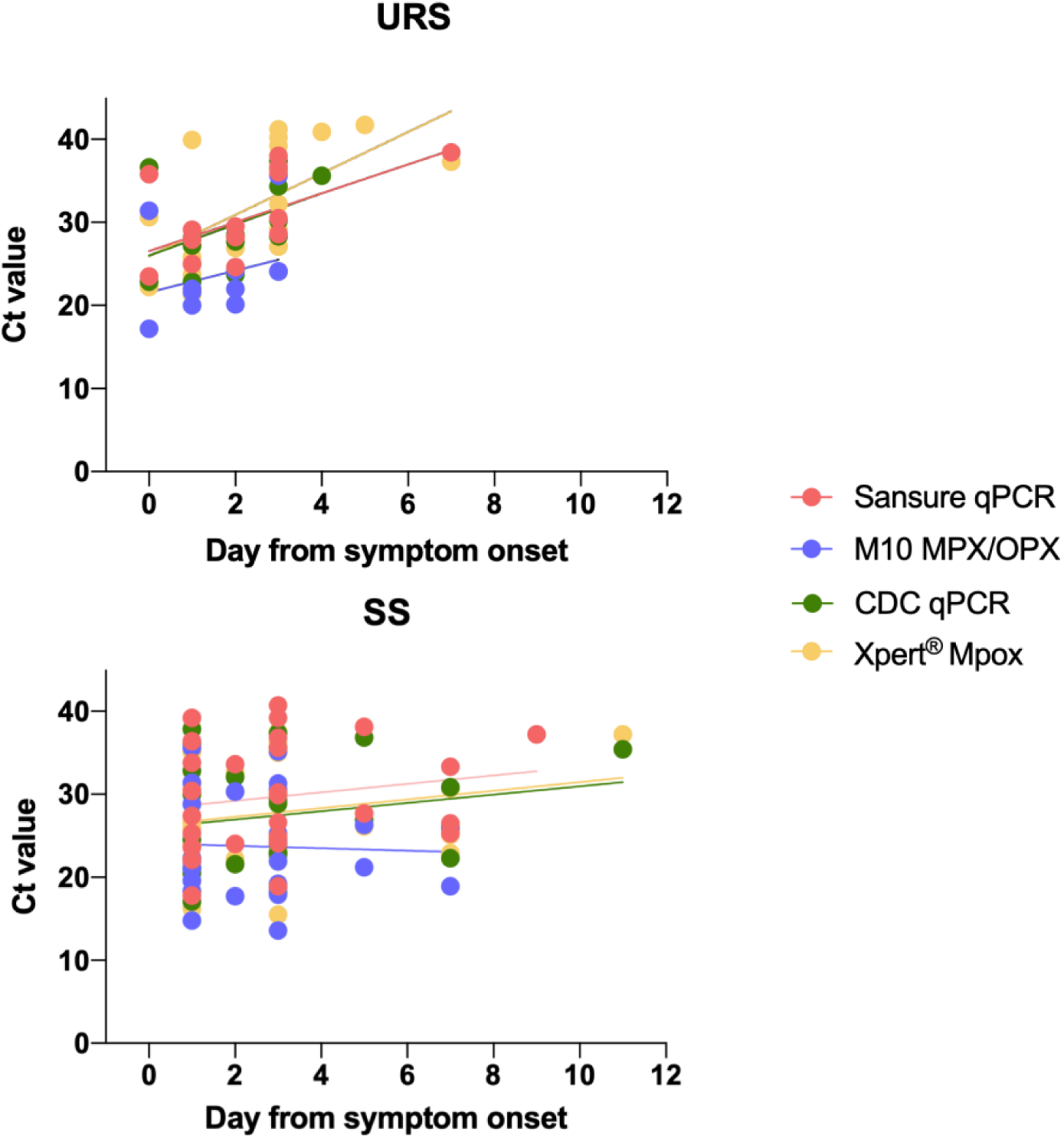
Plot of the Ct values of the four platforms by collection day from symptom onset using SS (n=30) and URS (n=23) from MPXV positive patients. Data points are individual clinical samples, with SS sampling from different lesions.

### Analytical evaluation

The LOD was 1.0×10^1^ pfu/mL for Xpert^®^ Mpox and M10 MPX/OPX, 1.0×10^2^ pfu/mL for Sansure qPCR and 1.0×10^3^ pfu/mL for the CDC qPCR (Figure 4). The approximated viral copy number of the LOD was calculated for all the assays and was at ≈1.31×10^2^ copies/mL for Xpert^®^ Mpox and M10 MPX/OPX, ≈1.3×10^3^ copies/mL for Sansure qPCR and ≈1.3×10^4^ copies/mL for CDC qPCR.

**Figure 4:**
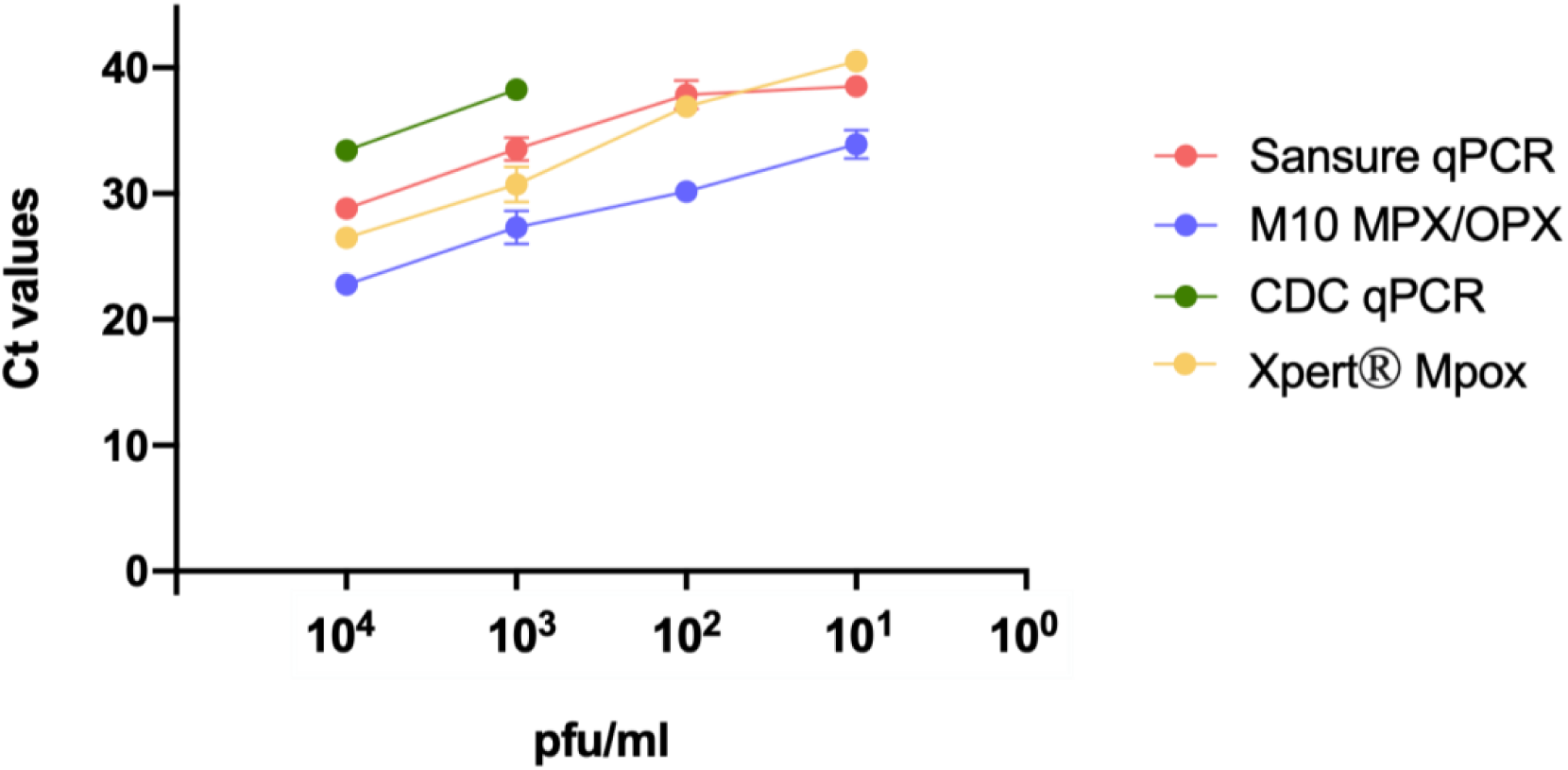
Relationship between Ct values and viral load using both qPCR reference assays and POC index NAATs.

## Discussion

The primary aim of this study was to evaluate the diagnostic performance of two POC NAAT, Xpert^®^ Mpox and M10 MPX/OPX. Rapid molecular diagnostic tests offer several advantages to laboratory-based PCR methods such as minimal sample processing, automated results readout and rapid availability of results to speed up clinical decision-making for timely management in outbreak situations, hence it is critical to assess their diagnostic accuracy.

The Xpert^®^ Mpox test is designed to be used with lesion swabs^30^. Previous studies have evaluated the accuracy for detection of MPXV in crusts and vesicular swabs samples in DRC showing a sensitivity of 98% and a specificity of 100% in both sample types^31^ and in oropharyngeal, lesions and anal swabs in Georgia, USA with a sensitivity of 100% and specificity of 83.3% to 90.9% depending on sample type^32^. The published data aligns with our results when using the platform with both lesion and upper-respiratory swabs. In this study, the Xpert^®^ Mpox detected MPXV DNA from clinical samples that were negative by the reference lab-based qPCR Sansure and CDC, suggesting greater sensitivity. The greater sensitivity of the Xpert^®^ Mpox compared to a reference lab-based MPXV PCR has also been observed elsewhere^32^. This could be due to the larger sample volume used in the Xpert^®^ Mpox (300 µl) compared to the volume used in the lab-based qPCR tests (2.5µl and 10µl of 50µl eluted DNA extracted from 200µl of UTM for CDC and Sansure respectively). The GeneXpert platform has been widely used for the detection of several infectious diseases, including SARS-CoV-2, *Mycobacterium tuberculosis* with rifampicin resistance, methicillin-resistant *Staphylococcus aureus*, and Ebola virus disease also showing high sensitivity and specificity^33–35^.

The M10 MPX/OPX can be used on different specimen types such as skin lesion material, whole blood, oropharyngeal swabs and plasma^36^. Compared to Xpert^®^ Mpox, sensitivity was lower despite of manufacturer claims of having the same analytical LOD (100 copies/mL) as the Sansure reference test. The M10 MPX/OPX assay is for research use and skin lesion material, whole blood, oropharyngeal swabs and plasm. In this study, the M10 MPX/OPX platform detected less MPXV positive samples than the other tests, suggesting lower sensitivity. A previous study evaluating the diagnostic accuracy of the M10 MPX/OPX test, showed lower sensitivity compared to the lab-based qPCR RealStar® OPX-1^37^, aligning with the results obtained in the present study. During the COVID-19 pandemic, SD Biosensor developed the M10 SARS-CoV-2, a molecular in vitro diagnostic assay able to detect SARS-CoV-2 viral RNA that also uses the M10 platform as the MPX/OPX assay^38^ with 100% sensitivity and 100% specificity^39,40^.

The WHO recommends the use of skin lesions for laboratory confirmation of MPXV infection whenever possible^15^. Our study indicates that URS can be used as a reliable alternative sample type to SS for patients sampled within the first 3 days of symptoms onset. This presents an advantage as the use of URS for POC testing in suspected cases can be used to diagnose Mpox in patients without typical skin lesions including those who may be in the prodromal phase of the disease when skin lesions have not appeared yet. The use of URS for Mpox diagnosis early in the disease can be particularly beneficial for monitoring contacts of positive cases for rapid detection, isolation and patient management. However, the use of SS for MPXV detection was more robust among samples collected from patients regardless of the time from symptom onset, except for M10 MPX/OPX that showed poor sensitivity in skin lesion swabs collected from patients more than 5 days after symptom onset. This provides key information for choosing the adequate sample type and tests, specifically for when patients present to the clinic several days after the disease onset. The Ct value of paired URS and SS showed no significant differences except for the Xpert^®^ Mpox platform. These results differ from previous studies where they observed that lesion samples presented a viral load 3 orders of magnitude higher than URS^41^ and suggest that URS testing offers no additional information for the diagnosis in individuals presenting skin lesions^41–43^. The lower sensitivity obtained in URS among these collected from patients more than 3 days after symptom onset can be attributed to viral clearance occurring earlier in the oropharynx sample than in skin lesions^44^.

Based on the target products profile (TPP) for tests used for mpox diagnosis within health care settings and laboratories published by the WHO, the minimal and optimal clinical sensitivity should be ≥ 95% and ≥ 97%, and minimal and optimal clinical specificity should be ≥ 97% compared to a reference molecular method^45^. The results obtained using the Xpert^®^ Mpox assay met the minimal clinical sensitivity using SS and optimal sensitivity using URS regardless of the qPCR used as reference method. In the case of the MPX/OPX assay, the minimal and optimal clinical sensitivity was met with SS when compared to Sansure and CDC qPCR reference tests however the sensitivity using URS did not fulfil the minimum clinical sensitivity regardless of the qPCR reference assay used. False positive results in the index tests have been attributed to lower sensitivity of the reference test compared to the index tests since all the “false positive” results were obtained from MPXV positive patients and both Xpert^®^ Mpox and M10 MPX/OPX had a lower LOD than the reference tests used in this study. This is of importance as reference lab-based qPCR tests such as the widely used CDC protocol may fail to diagnose true MPXV positive samples, and a composite reference standard should be determined.

The WHO recommends using laboratory-based nucleic acid amplification testing (NAAT) to confirm an MPXV infection^15^. The primers and probes used in the current MPXV generic qPCR test created by the CDC^21^ differ significantly due to genetic variations in >1000 available sequenced MPXV genomes impacting on the sensitivity and specificity of the test^46^. This could be a possible explanation of the higher LOD, and consequently lesser number of positive samples detected with the CDC qPCR compared with Sansure qPCR.

In this study we used frozen samples due to low prevalence causing difficulties for fresh samples and prospective evaluation. However, the IFU of both index tests indicate they can detect MPXV in frozen samples as well as samples stored at 4°C and room temperature^30^. The lack of negative skin lesion swab specimens is a limitation of the study as we could not calculate specificity using this sample type.

In conclusion, the Xpert^®^ Mpox demonstrated the greatest diagnostic accuracy for POC testing and the use of URS as alternative sample type to skin lesions have been shown to perform well in samples collected within 3 days from onset symptoms. This study adds important insights on diagnostics of Mpox.

## Author contributions

The study was conceived and designed by AICA, JD and DE. Laboratory work was conducted by AS, JP, YH, DW, NK, CTW and ARR, with supervision by AICA and TE. Data collections were conducted by AS, JP, YH, HH, NK, and SG. Data analysis and interpretation were conducted by ARR and AICA. The initial manuscript was prepared by ARR and AICA. Funding was acquired by AICA, the CONDOR steering group and ISARIC CPP investigators. Oversight of participant recruitment was performed by AICA, the CONDOR steering group and ISARIC CPP investigators. All authors edited and approved the final manuscript.

## Declarations of interests

DE had no role on the data collection, analysis and interpretation. The other authors have no interests to declare. The views expressed are those of the authors and not necessarily those of the funding bodies.

## Data Availability

All data produced in the present study are available upon reasonable request to the authors

## Acknowledgements

We acknowledge the participants for volunteering for this study. We thank all the CRN, research nurses and clinical research fellows for supporting us with the sample collection and recruitment. We also would like to extend our acknowledgements to the wider FIND team involved in this study, specifically to Audrey Albertini, Juvenal Nkeramahame, Michael Otieno and Ryan Ruiz.

*CONDOR steering group*: A. Joy Allen, Julian Braybrook, Peter Buckle, Eloise Cook, Paul Dark, Kerrie Davis, Gail Hayward, Adam Gordon, Anna Halstead, Charlotte Harden, Colette Inkson, Naoko Jones, William Jones, Dan Lasserson, Joseph Lee, Clare Lendrem, Andrew Lewington, Mary Logan, Massimo Micocci, Brian Nicholson, Rafael Perera-Salazar, Graham Prestwich, D. Ashley Price, Charles Reynard, Beverley Riley, John Simpson, Valerie Tate, Philip Turner, MarkWilcox, Melody Zhifang.

*ISARIC CCP investigators*: Dr Mike Beadsworth, Dr Ingeborg Welters, Dr Lance Turtle, Dr Jane Minton, Karl Ward, Dr Elinor Moore, Dr Elaine Hardy, Dr Mark Nelson, Dr Jane Minton, Karl Ward, Dr David Brealey, Dr Ashley Price, Dr Brian Angus, Dr Graham Cooke and Dr Oliver Koch.

## Funding

This work was funded as part of FIND’s work as coconvener of the diagnostics pillar of the Pandemic Threats Programme. ISARIC4 was funded from the National Institute for Health Research [award CO-CIN-01], the Medical Research Council [grant MC_PC_19059] and by Liverpool Pandemic Institute and the National Institute for Health Research Health Protection Research Unit (NIHR HPRU) in Emerging and Zoonotic Infections at University of Liverpool in partnership with UK Health Security Agency (UK-HSA), in collaboration with Liverpool School of Tropical Medicine and the University of Oxford [NIHR award 200907], Wellcome Trust and Department for International Development [215091/Z/18/Z], and the Bill and Melinda Gates Foundation [OPP1209135], and Liverpool Experimental Cancer Medicine Centre for providing infrastructure support for this research (Grant Reference: C18616/A25153).

The FALCON study was funded by the National Institute for Health Research, Asthma United Kingdom, and the British Lung Foundation. This work is partially funded by the National Institute for Health Research (NIHR) Health Protection Research Unit in Emerging and Zoonotic Infections (200907), a partnership between the United Kingdom Health Security Agency (UKHSA), The University of Liverpool, The University of Oxford, and LSTM.

## Notes

### Competing Interest Statement

The authors have declared no competing interest.

### Funding Statement

This work was funded as part of FIND work as coconvener of the diagnostics pillar of the Pandemic Threats Programme. ISARIC4 was funded from the National Institute for Health Research [award CO-CIN-01], the Medical Research Council [grant MC_PC_19059] and by Liverpool Pandemic Institute and the National Institute for Health Research Health Protection Research Unit (NIHR HPRU) in Emerging and Zoonotic Infections at University of Liverpool in partnership with UK Health Security Agency (UK-HSA), in collaboration with Liverpool School of Tropical Medicine and the University of Oxford [NIHR award 200907], Wellcome Trust and Department for International Development [215091/Z/18/Z], and the Bill and Melinda Gates Foundation [OPP1209135], and Liverpool Experimental Cancer Medicine Centre for providing infrastructure support for this research (Grant Reference: C18616/A25153). The FALCON study was funded by the National Institute for Health Research, Asthma United Kingdom, and the British Lung Foundation. This work is partially funded by the National Institute for Health Research (NIHR) Health Protection Research Unit in Emerging and Zoonotic Infections (200907), a partnership between the United Kingdom Health Security Agency (UKHSA), The University of Liverpool, The University of Oxford, and LSTM.

### Author Declarations

Patients were consented under the WHO ISARIC4 Comprehensive Clinical Characterisation Collaboration Protocol for severe emerging infections [ISRCTN66726260]20, ethical approval was obtained from the National Research Ethics Service and the Health Research Authority (IRAS ID:126600, REC 13/SC/0149). The COVID-19 samples were collected under the Facilitating AcceLerated Clinical validation Of Novel diagnostics for COVID-1919,23 and ethical approval was obtained from the National Research Ethics Service and the Health Research Authority (IRAS ID:28422, REC: 20/WA/0169).

